# Baseline Statin Exposure and Incident Acute Myocardial Infarction in Adults Aged ≥75 Years Without Prior Cardiovascular Disease: A Population-Based Cohort Study

**DOI:** 10.64898/2026.08.21.26361002

**Authors:** J. Cárdenas-Valladolid, O. Alonso-del Cura, M. Beneito-Dura, F.J. Somolinos-Simon, J. Mostaza, C. Lahoz, F.J. San Andrés-Rebollo, P. Vich-Pérez, AI. González-González, M.A. Salinero-Fort, the HealthData@MAD-R&I® Working Group

**Affiliations:** Dirección General de Investigación y Docencia, Consejería de Sanidad de la Comunidad de Madrid, Spain; Frailty, Multimorbidity Patterns and Mortality in the Elderly Population Residing in the Community, Hospital La Paz Institute for Health Research (IdiPAZ), Madrid, Spain; Universidad Alfonso X el Sabio, Madrid, Spain; Fundación para la Investigación e Innovación Biosanitaria de Atención Primaria (FIIBAP), Madrid, Spain; Unidad de Lípidos, Hospital La Paz-Carlos III, Madrid, Spain; Centro de Salud Las Calesas, Madrid, Spain; Centro de Salud Los Alpes, Madrid, Spain; Network for Research on Chronic Diseases, Primary Care, and Health Promotion (RICAPPS), Madrid, Spain; Instituto de Investigación Sanitaria Gregorio Marañón, Madrid, Spain

**Keywords:** Statins, primary prevention, acute myocardial infarction, aged ≥75 years, landmark analysis, competing risks, propensity-score matching, real-world data

## Abstract

**Background:** Adults aged ≥75 years represent a rapidly growing population at risk of acute myocardial infarction (AMI), yet they remain markedly underrepresented in statin trials for primary prevention. The limited evidence base, together with multimorbidity, functional heterogeneity, and competing mortality risks, has contributed to uncertainty regarding the potential role of statins in very old adults. This study evaluated the association between baseline statin exposure and incident AMI among community-dwelling adults aged ≥75 years without prior cardiovascular disease.

**Methods:** We conducted a retrospective population-based cohort study using linked primary-care, hospital, laboratory, and pharmacy dispensing data from the Community of Madrid. Statin exposure was ascertained during a 24-month exposure-assessment period from 1 January 2018 to 31 December 2019 and classified at a landmark date of 1 January 2020, when outcome follow-up began. Participants were classified as exposed if they had received at least two statin dispensations during the exposure-assessment period and had no record of prior lipid-lowering therapy before 2018. Individuals with prior cardiovascular disease, type 1 diabetes, cancer, dementia, or advanced chronic kidney disease were excluded. Missing data were addressed using multiple imputation. The association between baseline statin exposure and incident AMI was estimated using multivariable Cox proportional hazards regression. Propensity-score matching and Fine–Gray competing-risk regression, with all-cause mortality as the competing event, were performed as sensitivity analyses.

**Results:** Among 174,014 individuals included in the final cohort, 32,698 (18.8%) met the criteria for baseline statin exposure. The mean age was 82.5 years. During a median follow-up of 5 years, AMI occurred in 533 (1.63%) statin-exposed individuals and 2722 (1.93%) non-exposed individuals (p=0.0003). The observed absolute risk difference was 0.30 percentage points (95% CI, 0.14–0.45), corresponding to an estimated observational number needed to treat of 338 over 5 years (95% CI, 222–708). In the fully adjusted Cox model, baseline statin exposure was associated with a lower risk of incident AMI (HR, 0.805; 95% CI, 0.731–0.887). In the full-cohort Fine–Gray model accounting for competing mortality, baseline statin exposure remained associated with a lower cumulative incidence of AMI (sHR, 0.823; 95% CI, 0.748–0.905). After propensity-score matching, the association remained in the competing-risk analysis (subdistribution HR, 0.852; 95% CI, 0.738–0.983).

**Conclusions:** In this large population-based cohort of adults aged ≥75 years without prior cardiovascular disease, baseline statin exposure was associated with a lower incidence of AMI across several analytical approaches. The observed absolute risk difference was modest, and the findings should be interpreted in light of the observational design, residual confounding, and the potential for selection related to survival to the landmark date. Further randomized evidence is needed to determine whether this association reflects a causal effect of statin therapy in very old adults.

## Introduction

Older adults represent the fastest-growing segment of the population at risk for cardiovascular events. In Spain, the REGICOR study reported that the cumulative incidence of acute myocardial infarction (AMI) in individuals older than 74 years ranges from 1111 to 2306 per 100,000 in men and from 576 to 1384 per 100,000 in women, depending on age group (1). Approximately 60% of patients hospitalized for AMI are aged ≥75 years, and nearly 65% of AMI-related deaths occur in this age group; yet adults over 75 rarely represent more than 7% of participants in clinical studies (2). This persistent underrepresentation limits the generalizability of existing evidence to the very old.

In addition, older adults experience substantial functional and psychological consequences after an AMI. A marked decline in independence has been documented after hospital discharge, particularly following ST-elevation myocardial infarction (STEMI) (3), along with increased anxiety and depressive symptoms (4). These complications hinder recovery, reduce adherence to long-term therapies, and amplify the long-term impact of a first coronary event. Such vulnerability underscores the clinical relevance of preventing AMI in very old adults, for whom the consequences extend far beyond short-term mortality and profoundly affect quality of life, autonomy, and subsequent healthcare needs.

Despite the proven efficacy of statins in randomized trials among middle-aged populations, evidence supporting their use for primary prevention beyond age 75 remains limited and controversial. Randomized trials have underrepresented this age group, and observational studies are challenged by heterogeneity in comorbidity burden, frailty, and competing mortality risks. According to the START criteria (5), statins and antiplatelet agents are among the most frequently omitted recommended therapies in older adults with diabetes or cardiovascular disease (6), further highlighting gaps in preventive care. In this context, focusing on AMI, a direct, mechanistically linked outcome, offers a more specific assessment of the association between statins and AMI risk, minimizing confounding from non-cardiovascular causes of death.

This study therefore aimed to evaluate the association between baseline statin exposure and the incidence of AMI over five years among community-dwelling adults aged ≥75 years without prior cardiovascular disease, type 1 diabetes, cancer, dementia, advanced chronic kidney disease, or previous cardiovascular medication exposure.

## Material and Methods

This retrospective population-based cohort study was conducted within HealthData@MAD-R&I^®^, a regional health data space initiative that supports the secure and responsible secondary use of routinely collected health information for research, innovation, and evidence-informed policymaking. The initiative operates under robust European and national data governance frameworks, ensuring compliance with ethical, legal, and technical standards for data protection and interoperability. One of its real-world use cases examines the potential association between baseline statin exposure and incident AMI.

We identified all community-dwelling individuals aged 75 years or older who were residing in the Community of Madrid on January 1, 2020. This date was defined as the landmark date and the beginning of outcome follow-up. A total of 636,281 individuals were initially identified.

The study used a fixed baseline exposure classification. Statin exposure was determined from pharmacy dispensing records during the 24-month exposure-assessment period (1 January 2018 to 31 December 2019). Eligibility and exposure status were defined at the landmark date, after which participants were followed prospectively for incident acute myocardial infarction.

Participants were classified as exposed to statins if they had received at least two statin dispensations during the exposure-assessment period and had no record of prior lipid-lowering therapy before 1 January 2018. Participants meeting this criterion received a median of 21 statin dispensations during the exposure-assessment period, with a range of 2 to 24 prescriptions. Participants were analyzed according to their baseline exposure classification, irrespective of subsequent treatment discontinuation, treatment interruptions, initiation of statins among those classified as unexposed at baseline, or changes in treatment intensity.

Because eligibility was assessed at the landmark date, the study population was necessarily restricted to individuals who remained alive, registered, and observable until 1 January 2020. This design ensured that exposure status was fully defined before outcome follow-up began.

### Exclusions

Patients with previous cardiovascular disease, including peripheral artery disease, ischemic or hemorrhagic stroke, heart failure, or coronary heart disease such as angina or myocardial infarction, or those receiving cardiac therapy drugs (ATC class C01) were excluded to minimize confounding by indication, as these medications are typically prescribed for established cardiovascular disease. This criterion ensured that the analysis focused on individuals without prior cardiac conditions, allowing a more accurate assessment of the association between statin use and incident myocardial infarction.

We also excluded individuals with any exposure to lipid-lowering medications before 2018. In addition, participants with type 1 diabetes, cancer, dementia, or advanced chronic kidney disease were excluded to minimize confounding from conditions that markedly alter metabolic or hemodynamic profiles.

The study flowchart is shown in Figure 1. The final cohort comprised 174,014 individuals, of whom 18.8% were exposed to statins (n = 32,698).

**Figure 1.**
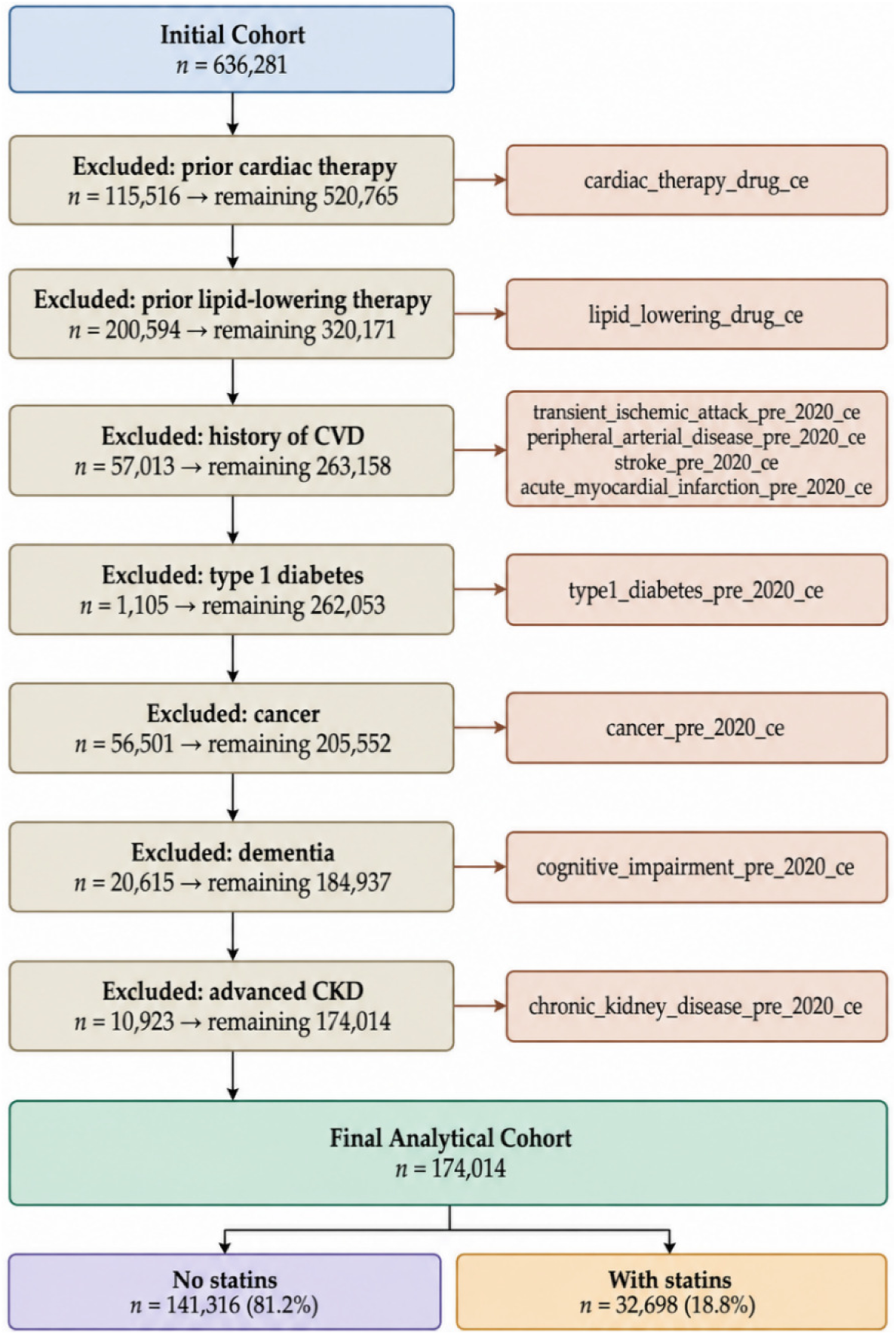
Flowchart.

### Data Sources

Data were obtained from the same sources used to construct the Aged-Madrid cohort (7). Primary care electronic medical records (EMRs) provided information on diagnoses, cardiovascular risk factors, lifestyle characteristics, functional status, and laboratory measurements, and have been previously validated for major chronic conditions (8) (9). Hospital discharge records (CMBD) were used to ascertain diagnoses, including cardiovascular diseases, cancer, and renal conditions. Laboratory information systems supplied biochemical measurements, and pharmacy dispensing records from the Official College of Pharmacists of Madrid captured medication exposure.

### Study Variables

Baseline comorbidities were identified from diagnoses recorded before the landmark date using ICD-9, ICD-10, and ICPC-2 codes. The assessed conditions included hypertension, dyslipidemia, obesity, type 2 diabetes, atrial fibrillation, ischemic heart disease, stroke, heart failure, peripheral arterial disease, cancer, dementia, tobacco use, chronic obstructive pulmonary disease, and chronic kidney disease.

Socioeconomic context was assessed using the MEDEA deprivation index (10), an area-level indicator derived from small geographic units. The index incorporates neighborhood characteristics such as unemployment, low educational attainment, manual occupations, and temporary employment. Scores are standardized, with higher values indicating greater deprivation.

Clinical and functional variables included body mass index (BMI), waist circumference, systolic and diastolic blood pressure, number of blood pressure measurements, Barthel Index, Lawton-Brody instrumental activities of daily living index, LDL cholesterol, number of LDL cholesterol measurements, triglycerides, fasting plasma glucose, microalbuminuria, and estimated glomerular filtration rate calculated using the CKD-EPI equation. For each variable, the measurement used for analysis was the prespecified value obtained during the exposure-assessment period.

Medication exposure was identified from pharmacy dispensing records and classified according to the following Anatomical Therapeutic Chemical (ATC) codes: C01, C10AA, C10AB, C10AC, C10AD, C10AX, A10BH, A10BJ, A10BK, B01AC, B01AE, B01AF, B01AC06, C07A, C07B, C07C, C07D, C08C, C08D, C09A, C09B, C09C, and C09D.

The primary outcome was the first incident acute myocardial infarction (AMI) during follow-up. AMI was identified using primary care electronic medical records and hospital discharge data, based on validated ICD-10 (I21–I22) and ICD-9 codes (410) and the corresponding ICPC-2 code (K75). The coding algorithm used to identify AMI has been previously validated in our setting using primary-care clinical records (9).

Mortality was identified through the CIBELES regional healthcare information system rather than through linkage with official mortality registries. Information on the specific cause of death was not available when death occurred.

### Data Processing and Quality Control

#### Data Curation

Before construction of the analytical variables, systematic quality-control procedures were performed on the raw dataset. Missingness was quantified for all variables and missing-data patterns were examined. Data types were checked and corrected when variables were stored in formats inconsistent with their expected definition (e.g., dates stored as strings). Duplicate records were removed, retaining one unique observation per individual.

Records after 31 December 2024 were excluded. Clinical and laboratory variables underwent a standardised cleaning and normalisation process. Raw values were harmonised to predefined units of measurement, with conversions applied where necessary. Plausibility filters were applied to exclude implausible values. Intra-patient outliers were identified using intra-patient outlier detection methods applied to each patient’s individual measurement distribution, using either robust or standard z-scores depending on the variable. Additionally, diagnostic codes from ICPC-2 were reviewed for consistency between codes and descriptors, and recoding was performed where inconsistencies were identified. Medication exposures were constructed from pharmacy dispensing records using prespecified ATC codes and exposure definitions.

#### Missing Data

Several variables contained missing values, with the proportion of missingness ranging from 4.7% to 93.5%. The proportion of missingness for each variable is presented in Supplementary Table S1. As the imputation of variables with extremely high rates of missingness can introduce substantial uncertainty, a pre-specified threshold of 70% missingness was established. On this basis, three variables were removed from the analytical pipeline: waist circumference (93.5%), microalbuminuria (72.6%), and the Lawton–Brody IADL scale (71.2%). All remaining variables fell below this threshold and were retained for analysis.

The overall pattern of missing data was compatible with a Missing at Random (MAR) mechanism conditional on observed covariates. Therefore, multiple imputations by chained equations (MICE) were applied, implemented via the miceforest package in Python, which applies predictive mean matching (PMM) to impute missing values from observed values in clinically similar patients. Twenty imputed datasets were generated using 10 iterations each with the miceforest package (version 6.0.5) in Python. Predictive mean matching was performed using five candidate donors and the default normal mean-matching strategy, with a fixed random seed of 42. The outcome variable and follow-up time were included in the imputation model. Results from Cox regression models fitted to each imputed dataset were combined using Rubin’s rules. Comparisons between the original distribution of each variable and the corresponding distributions across the 20 imputed datasets are presented in Supplementary Figures S3 and S4.

### Statistical Analysis

Continuous variables were summarized as means with standard deviations (SDs) or medians with interquartile ranges (IQRs), as appropriate, and categorical variables as counts and percentages. Between-group comparisons were performed using the Student’s t-test, analysis of variance, Mann–Whitney U test, Kruskal–Wallis test, chi-square test, or Fisher’s exact test, as appropriate.

To minimize bias related to changes in treatment exposure during follow-up, statin use was defined according to fixed baseline statin exposure status. Baseline comparability was assessed using standardized mean differences (SMDs), which are independent of sample size.

Incidence rates of AMI were calculated per 1000 person-years, stratified by sex. Ninety-five percent confidence intervals were obtained using exact Poisson methods.

The association between baseline statin exposure and incident AMI was estimated using multivariable Cox proportional hazards regression. The primary analysis followed a fixed baseline exposure strategy: participants remained classified according to their exposure status at the landmark date, regardless of subsequent changes in treatment.

Covariates were selected a priori based on clinical relevance and their potential association with statin exposure and AMI risk. The prespecified model included age, sex, Barthel Index, tobacco use, MEDEA deprivation index, body mass index, systolic and diastolic blood pressure, estimated glomerular filtration rate, hypertension, type 2 diabetes, atrial fibrillation, triglycerides, and concomitant use of aspirin, calcium-channel blockers, ACE inhibitors, angiotensin II receptor blockers, and DPP-4 inhibitors.

Follow-up began on 1 January 2020 and continued until the earliest occurrence of incident AMI, death, permanent removal from the CIBELES registry, or 31 December 2024. Death was treated as a competing event in the competing-risk analysis, whereas permanent removal from the registry was handled as a censoring event.

The proportional hazards assumption was assessed using Schoenfeld residuals and global tests of proportionality. Functional form and model fit were evaluated using Martingale and deviance residuals. Influential observations were assessed using dfbeta statistics, and multicollinearity was evaluated using variance inflation factors.

### Propensity-Score Analysis

As a sensitivity analysis and complementary approach to confounding control, propensity scores were estimated using logistic regression. The propensity-score model included the full set of baseline covariates considered in the initial analysis.

One-to-one nearest-neighbor matching without replacement was performed using a caliper width of 0.20 of the standard deviation of the logit of the propensity score. Covariate balance before and after matching was assessed using absolute standardized mean differences, with an absolute SMD below 0.10 considered indicative of adequate balance. Balance was also assessed graphically using a Love plot.

The matched cohort was subsequently analyzed using Fine–Gray subdistribution hazard regression, with all-cause mortality treated as the competing event. The post-matching Fine–Gray model was additionally adjusted using the full set of baseline covariates included in the initial model, without backward variable selection, to account for residual imbalance and improve precision. Estimates from this analysis were interpreted as associations with the cumulative incidence of AMI in the presence of competing mortality.

### Software and Statistical Significance

All statistical analyses were performed using Python version 3.10.16 and the corresponding libraries (psmpy 0.3.16, miceforest 6.0.5 and cmprsk 1.1.1), together with RStudio version 4.1.3 (survival, cmprsk). All tests were two-sided, and a p-value <0.05 was considered statistically significant. Given the observational nature of the study, the resulting estimates were interpreted as associations rather than causal treatment effects.

## Results

### Baseline Characteristics

A total of 174,014 individuals aged ≥75 years were included, of whom 18.8% (n = 32,698) were exposed to statins during 2018–2019. Compared with non-users, statin users were slightly younger (81.6 vs. 82.7 years; p < 0.001; SMD = –0.207) and lived in slightly more deprived areas, as reflected by higher MEDEA scores (–0.7 vs. –0.8; p < 0.001; SMD = 0.093). Female sex was slightly more frequent among statin users (67.4% vs. 66.6%; p = 0.007; SMD= 0.017). Statin users exhibited a significantly higher prevalence of dyslipidemia, as expected, and showed higher frequencies of hypertension, obesity, atrial fibrillation, COPD, tobacco use, and type 2 diabetes, with standardized mean differences ranging from 0.10 to 0.38 (Table 1). These differences reflect a clinically meaningful clustering of cardiometabolic risk factors among individuals receiving statin therapy.

**Table 1.** Baseline characteristics of the study population by statin exposure (2018–2019)

|  | Missing | Overall<br>(n=174,014) | No statins<br>(n=141,316) | Statins<br>(n=32,698) | SMD | p-value |
| --- | --- | --- | --- | --- | --- | --- |
| **Demographics** |  |  |  |  |  |  |
| Age, mean (SD) | 0 | 82.5 (5.6) | 82.7 (5.7) | 81.6 (4.9) | -0.207 | <0.001 |
| Deprivation index, mean (SD) | 8,154 | -0.8 (0.9) | -0.8 (0.9) | -0.7 (0.8) | 0.093 | <0.001 |
| Sex, n (%) | 0 |  |  |  | 0.017 | 0.007 |
| *Female* |  | 116,172 (66.8) | 94,134 (66.6) | 22,038 (67.4) |  |  |
| *Male* |  | 57,842 (33.2) | 47,182 (33.4) | 10,660 (32.6) |  |  |
| **Comorbidities** |  |  |  |  |  |  |
| Hypertension, n (%) | 0 | 121,591 (69.9) | 96,557 (68.3) | 25,034 (76.6) | 0.185 | <0.001 |
| Dyslipidemia, n (%) | 0 | 68,703 (39.5) | 41,139 (29.1) | 27,564 (84.3) | **1.341** | <0.001 |
| Obesity, n (%) | 0 | 46,184 (26.5) | 35,372 (25.0) | 10,812 (33.1) | 0.178 | <0.001 |
| Atrial fibrillation, n (%) | 0 | 20,418 (11.7) | 15,682 (11.1) | 4,736 (14.5) | 0.102 | <0.001 |
| COPD, n (%) | 0 | 11,408 (6.6) | 9,024 (6.4) | 2,384 (7.3) | 0.036 | <0.001 |
| Tobacco use, n (%) | 0 | 8,964 (5.2) | 7,025 (5.0) | 1,939 (5.9) | 0.042 | <0.001 |
| Type 2 diabetes, n (%) | 0 | 31,539 (18.1) | 21,459 (15.2) | 10,080 (30.8) | 0.378 | <0.001 |
| **Functional status** |  |  |  |  |  |  |
| Barthel index category, n (%) | 88,905 |  |  |  | 0.215 | <0.001 |
| *Independent (0)* |  | 47,255 (27.2) | 36,402 (25.8) | 10,853 (33.2) |  |  |
| *Mild dependence (1)* |  | 10,572 (6.1) | 8,155 (5.8) | 2,417 (7.4) |  |  |
| *Moderate dependence (2)* |  | 20,804 (12.0) | 16,552 (11.7) | 4,252 (13.0) |  |  |
| *Severe dependence (3)* |  | 6,478 (3.7) | 5,578 (3.9) | 900 (2.8) |  |  |
| *Missing* |  | 88,905 (51.1) | 74,629 (52.8) | 14,276 (43.7) |  |  |
| Lawton IADL category, n (%) | 123,899 |  |  |  | 0.225 | <0.001 |
| *Independent (0)* |  | 36,018 (20.7) | 26,891 (19.0) | 9,127 (27.9) |  |  |
| *Mild limitation (1)* |  | 7,172 (4.1) | 5,584 (4.0) | 1,588 (4.9) |  |  |
| *Moderate limitation (2)* |  | 3,691 (2.1) | 3,020 (2.1) | 671 (2.1) |  |  |
| *Severe limitation (3)* |  | 3,234 (1.9) | 2,795 (2.0) | 439 (1.3) |  |  |

| Missing |  | 123,899 (71.2) | 103,026 (72.9) | 20,873 (63.8) |  |  |
| --- | --- | --- | --- | --- | --- | --- |
| **Clinical measurements** |  |  |  |  |  |  |
| BMI, mean (SD) | 81,066 | 28.1 (4.8) | 28.0 (4.8) | 28.5 (4.8) | 0.109 | <0.001 |
| Waist circumference, mean (SD) | 162,641 | 102.6 (12.6) | 102.6 (12.7) | 102.8 (12.4) | 0.016 | 0.442 |
| Systolic BP, mean (SD) | 45,988 | 134.7 (17.4) | 134.8 (17.4) | 134.2 (17.2) | -0.035 | <0.001 |
| Systolic BP measurements, mean (SD) | 45,988 | 9.6 (16.1) | 9.5 (16.3) | 10.0 (15.6) | 0.028 | <0.001 |
| Diastolic BP, mean (SD) | 46,085 | 73.7 (10.1) | 73.8 (10.2) | 73.1 (9.9) | -0.071 | <0.001 |
| eGFR CKD-EPI, mean (SD) | 83,570 | 71.9 (13.6) | 71.9 (13.5) | 71.7 (13.8) | -0.020 | 0.013 |
| LDL cholesterol, mean (SD) | 61,700 | 112.8 (29.0) | 116.1 (27.0) | 102.0 (32.6) | -0.468 | <0.001 |
| LDL measurements, mean (SD) | 61,700 | 1.9 (1.0) | 1.8 (1.0) | 2.1 (1.1) | 0.300 | <0.001 |
| Triglycerides, mean (SD) | 56,490 | 110.8 (49.4) | 108.4 (47.4) | 119.0 (54.7) | 0.207 | <0.001 |
| Plasma glucose, mean (SD) | 54,386 | 98.8 (24.0) | 97.4 (22.7) | 103.9 (27.3) | 0.261 | <0.001 |
| Microalbuminuria, mean (SD) | 126,258 | 14.5 (58.2) | 13.5 (47.2) | 17.3 (82.7) | 0.056 | <0.001 |
| **Medications use** |  |  |  |  |  |  |
| ACE inhibitor or ARB, n (%) | 0 | 87,857 (50.5) | 66,928 (47.4) | 20,929 (64.0) | 0.340 | <0.001 |
| Antiplatelet agents, n (%) | 0 | 23,089 (13.3) | 14,461 (10.2) | 8,628 (26.4) | 0.427 | <0.001 |
| Aspirin, n (%) | 0 | 21,589 (12.4) | 13,770 (9.7) | 7,819 (23.9) | 0.386 | <0.001 |
| Beta-blockers, n (%) | 0 | 26,031 (15.0) | 18,337 (13.0) | 7,694 (23.5) | 0.276 | <0.001 |
| Calcium channel blockers, n (%) | 0 | 26,732 (15.4) | 19,840 (14.0) | 6,892 (21.1) | 0.186 | <0.001 |
| DPP4 inhibitors, n (%) | 0 | 9,277 (5.3) | 5,533 (3.9) | 3,744 (11.5) | 0.286 | <0.001 |
| SGLT2 inhibitors, n (%) | 0 | 1,588 (0.9) | 744 (0.5) | 844 (2.6) | 0.167 | <0.001 |
| GLP-1 receptor agonists, n (%) | 0 | 515 (0.3) | 170 (0.1) | 345 (1.1) | 0.123 | <0.001 |

Values are mean (SD) for continuous variables and n (%) for categorical variables. SMD: standardized mean difference. Missing: number of patients with missing values. BP: blood pressure; BMI: body mass index; eGFR: estimated glomerular filtration rate; CKD-EPI: Chronic Kidney Disease Epidemiology Collaboration; ACE: angiotensin-converting enzyme; ARB: angiotensin receptor blocker; SGLT2: sodium-glucose cotransporter-2; DPP4: dipeptidyl peptidase-4; GLP-1: glucagon-like peptide 1; IADL: instrumental activities of daily living. SMD >0.10 indicates potential imbalance; SMD >0.30 shown in red.

Functional status also differed between groups. Based on the available data, statin-exposed participants were more frequently classified as independent or mildly dependent than non-exposed participants. The standardized mean differences were 0.215 for the Barthel Index and 0.225 for the Lawton–Brody IADL scale. These comparisons should be interpreted in the context of the substantial missingness observed for both functional measures.

Clinical measurements revealed small to moderate differences. Statin users had slightly higher BMI (SMD= 0.109) and triglyceride levels (SMD= 0.207), lower LDL cholesterol (SMD= -0.468), and marginally lower systolic and diastolic blood pressure. Fasting plasma glucose was higher among statin users (SMD = 0.26), consistent with their higher prevalence of diabetes. Kidney function (eGFR) was similar between groups.

Medication patterns showed marked differences: statin users were substantially more likely to receive ACE inhibitors/ARBs, antiplatelet agents, beta-blockers, calcium-channel blockers, and glucose-lowering therapies (including DPP-4 and SGLT-2 inhibitors), with SMDs ranging from 0.17 to 0.43. These patterns reflect a higher burden of cardiometabolic disease and more intensive cardiovascular risk management among statin users (Table 1).

### Incidence of AMI

Overall, 1,553 men experienced an AMI during follow-up, corresponding to a cumulative incidence of 2.68% (1,553/57,842) and an incidence rate of 6.29 per 1,000 person-years. Among women, 1,702 AMI events were recorded, yielding a cumulative incidence of 1.47% (1,702/116,172) and an incidence rate of 3.35 per 1,000 person-years. Both differences were statistically significant (p < 0.001). The Kaplan–Meier curves for AMI-free survival showed a consistent separation between men and women throughout the 2020–2024 follow-up period. Women maintained a higher probability of remaining free from AMI at all time points, whereas the curve for men declined more steeply, indicating a greater cumulative incidence of events (Figure 2).

**Figure 2.**
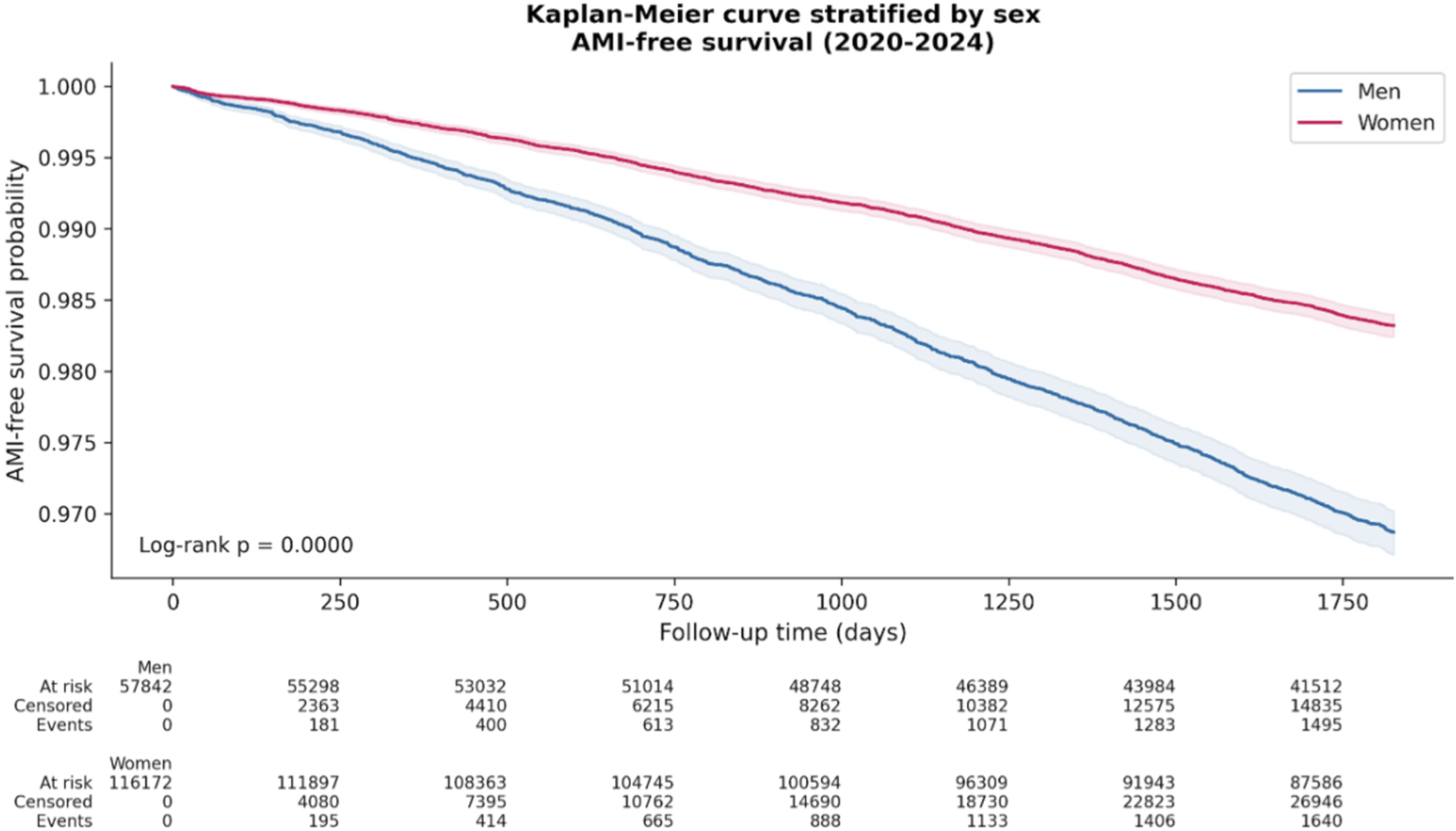
Kaplan-Meier curve stratified by sex.

The crude rate of AMI was 4.311 per 1000 person-years (95% CI, 4.164–4.461). When stratified by statin exposure, the rate was 3.673 per 1000 person-years (95% CI, 3.367–3.998) among statin users and 4.462 per 1000 person-years (95% CI, 4.296–4.633) among non-users. The rate ratio comparing statin users with non-users was 0.823 (95% CI, 0.749–0.903; p < 0.001).

Individuals treated with statins had a 5-year risk of AMI of 1.63% (533/32,698), which was significantly lower than the risk observed among those not receiving statins (1.93% (2722/141,316); p = 0.0003). Figure 3 shows the Kaplan–Meier curves, which began to diverge around day 500 of follow-up. From that point onward, statin users maintained consistently higher AMI-free survival probabilities than non-users. The separation between curves widened progressively, and the log-rank test confirmed a statistically significant difference in AMI-free survival (p < 0.001).

**Figure 3.**
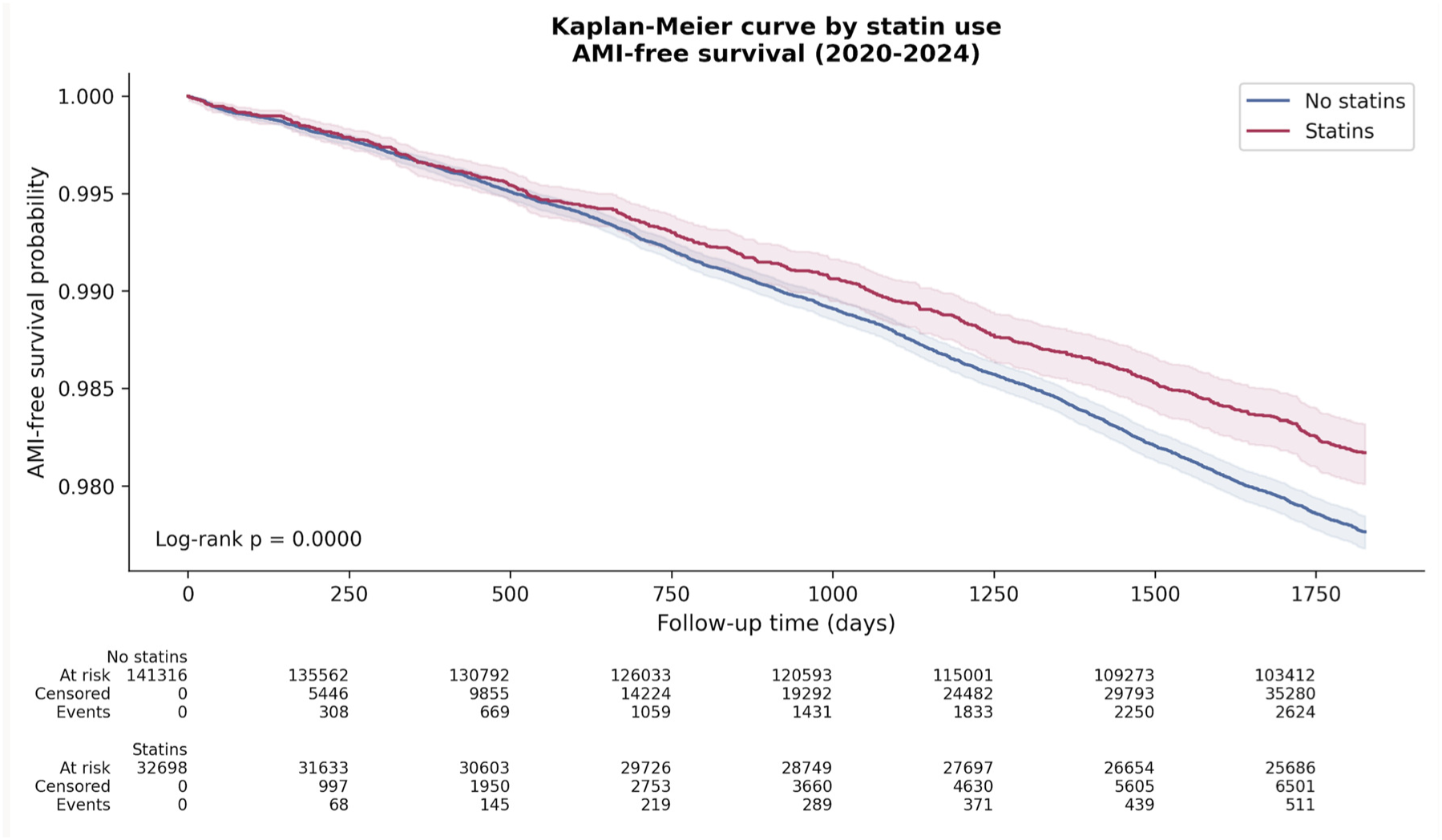
Kaplan-Meier curve stratified by statin use.

When examining absolute differences, statin therapy was associated with a modest variation in AMI risk. The observed absolute difference in AMI event proportions was 0.30% (95% CI 0.14–0.45). This corresponds to an estimated observational number needed to treat of 338 over 5 years (95% CI 222– 708), a metric that quantifies the absolute difference observed in this cohort.

A comparable separation of the curves was observed when stratifying by sex, as shown in Supplementary Figures S1 and S2.

### Multivariable analysis

In the primary Cox proportional hazards model (n = 174,014), adjusted statin exposure was associated with a significantly lower hazard of incident myocardial infarction (HR 0.805; 95% CI 0.731–0.887; p < 0.001) (Table 2).

**Table 2.**
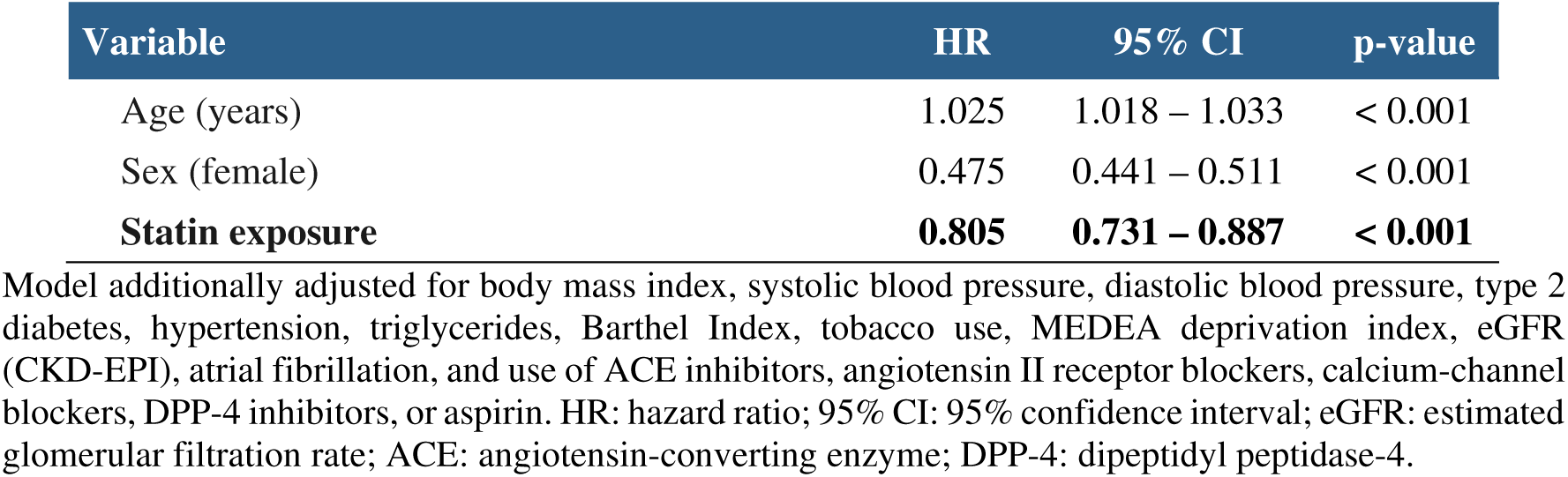
Cox regression analysis (n=174,014)

| Variable | HR | 95% CI | p-value |
| --- | --- | --- | --- |
| Age (years) | 1.025 | 1.018 – 1.033 | < 0.001 |
| Sex (female) | 0.475 | 0.441 – 0.511 | < 0.001 |
| **Statin exposure** | **0.805** | **0.731 – 0.887** | **< 0.001** |

Model additionally adjusted for body mass index, systolic blood pressure, diastolic blood pressure, type 2 diabetes, hypertension, triglycerides, Barthel Index, tobacco use, MEDEA deprivation index, eGFR (CKD-EPI), atrial fibrillation, and use of ACE inhibitors, angiotensin II receptor blockers, calcium-channel blockers, DPP-4 inhibitors, or aspirin. HR: hazard ratio; 95% CI: 95% confidence interval; eGFR: estimated glomerular filtration rate; ACE: angiotensin-converting enzyme; DPP-4: dipeptidyl peptidase-4.

The diagnostic plots indicated (Supplementary Figure S5) an overall good fit of the Cox proportional hazards model. The Martingale residuals showed no systematic pattern over follow-up time, with values symmetrically distributed around zero and without evidence of non-linearity or time-dependent misspecification. This supports the adequacy of the functional form of the continuous covariates included in the model.

The deviance residuals were similarly well-behaved. Most observations clustered within the ±2 range, with only a small number of points exceeding these thresholds, suggesting the absence of influential outliers or poorly fitted observations. The symmetric distribution around zero further indicates that the model captured the underlying hazard structure appropriately.

Taken together, these diagnostics support the robustness of the final Cox model and do not suggest violations that would compromise the validity of the estimated associations.

Given that 19.8% of participants died before experiencing myocardial infarction, mortality constituted a relevant competing event. Therefore, we performed a Fine–Gray competing-risk analysis (Table 3). In this model, statin exposure remained associated with a lower subdistribution hazard of AMI (sHR 0.823; 95% CI 0.748–0.905; p = 0.0001), indicating that the observed association persisted even after accounting for the non-negligible competing risk of death. The consistency between the Cox and Fine– Gray estimates reinforces the robustness of the findings.

**Table 3.** Fine–Gray competing-risk model for incident acute myocardial infarction, with mortality as the competing event.

| Variable | sHR | 95% CI | p-value |
| --- | --- | --- | --- |
| Age (years) | 1.009 | 1.001 – 1.017 | 0.0213 |
| Sex (women) | 0.509 | 0.473 – 0.547 | 0.0001 |
| **Statin exposure** | **0.823** | **0.748 – 0.905** | **0.0001** |

Model additionally adjusted for body mass index, systolic blood pressure, diastolic blood pressure, type 2 diabetes, hypertension, triglycerides, Barthel Index, tobacco use, MEDEA deprivation index, eGFR (CKD-EPI), atrial fibrillation, and use of ACE inhibitors, angiotensin II receptor blockers, calcium-channel blockers, DPP-4 inhibitors, or aspirin. sHR: subdistribution hazard ratio; 95% CI: 95% confidence interval; eGFR: estimated glomerular filtration rate; ACE: angiotensin-converting enzyme; DPP-4: dipeptidyl peptidase-4.

### Sensitivity Analysis

#### Propensity Score Matching

Baseline characteristics differed substantially between statin users and non-users before matching, as shown in Table 4. Prior to PSM, statin users were younger, had higher BMI, and exhibited a markedly greater prevalence of cardiometabolic comorbidities, including hypertension, type 2 diabetes, dyslipidemia, and obesity, with several SMD values exceeding 0.20 and some surpassing 1.0, indicating pronounced imbalance. Laboratory parameters also differed, with statin users showing lower LDL cholesterol and higher plasma glucose and triglyceride levels. Use of concomitant cardiovascular medications was consistently more frequent among statin users, further reflecting baseline clinical differences between groups.

**Table 4.** Baseline characteristics before and after propensity score matching according to statin exposure (2018–2019)

| Characteristic | Statins<br>(before)<br>N= 32,698 | No Statins<br>(before)<br>N= 141,316 | SMD<br>(before) | Statins<br>(after)<br>N= 26,747 | No Statins<br>(after)<br>N= 26,747 | SMD<br>(after) |
| --- | --- | --- | --- | --- | --- | --- |
| Propensity score (distance) | 0.4609 | 0.1247 | 1.3081 | 0.3888 | 0.369 | 0.0771 |
| Age, years | 81.63 | 82.73 | -0.2241 | 81.80 | 81.92 | -0.0234 |
| Female sex, % | 67.4 | 66.61 | 0.0168 | 68.67 | 69.00 | -0.0092 |
| Deprivation index | -0.7091 | -0.7889 | 0.0943 | -0.7301 | -0.7365 | 0.0077 |
| BMI, Kg/m<sup>2</sup> | 27.71 | 26.85 | 0.1897 | 27.53 | 27.43 | 0.0020 |
| Systolic blood pressure, mmHg | 134.22 | 134.35 | -0.0077 | 134.25 | 134.29 | -0.0020 |
| Diastolic blood pressure, mmHg | 73.20 | 73.74 | -0.0538 | 73.40 | 73.35 | 0.0053 |
| eGFR (CKD-EPI), mL/min/1.73 m<sup>2</sup> | 71.69 | 72.28 | -0.0431 | 71.87 | 71.82 | 0.0036 |
| Plasma glucose, mg/dL | 103.20 | 95.93 | 0.2698 | 100.80 | 100.47 | 0.0122 |
| LDL cholesterol, mg/dL | 101.84 | 115.89 | -0.4398 | 107.29 | 110.47 | -0.0994 |
| Triglycerides, mg/dL | 118.42 | 106.32 | 0.2232 | 116.88 | 117.53 | -0.00119 |
| Hypertension, % | 76.56 | 68.33 | 0.1944 | 74.99 | 75.11 | -0.0027 |
| Atrial fibrillation, % | 14.48 | 11.1 | 0.0962 | 13.86 | 13.79 | -0.0018 |
| COPD, % | 7.29 | 6.39 | 0.0348 | 7.01 | 6.84 | 0.0068 |
| Tobacco use, % | 5.93 | 4.97 | 0.0406 | 5.73 | 5.56 | -0.0070 |
| Obesity, % | 33.07 | 25.03 | 0.1708 | 31.47 | 30.73 | 0.0157 |
| Dyslipidemia, % | 84.3 | 29.11 | 1.5169 | 80.89 | 81.34 | -0.0122 |
| Barthel category: independent, % | 60.45 | 57.81 | -0.0541 | 60.41 | 60.02 | 0.080 |
| Type 2 diabetes, % | 30.83 | 15.19 | 0.3387 | 25.67 | 25.13 | 0.0117 |
| Antiplatelet agents, % | 26.39 | 10.23 | 0.3665 | 20.35 | 19.73 | 0.0141 |
| Aspirin, % | 23.91 | 9.74 | 0.3322 | 18.76 | 18.44 | 0.0075 |
| Beta-blockers, % | 23.53 | 12.98 | 0.2488 | 20.32 | 20.11 | 0.0050 |
| Calcium channel blockers, % | 21.08 | 14.04 | 0.1726 | 19.16 | 18.59 | 0.0139 |
| ACE inhibitor or ARB, % | 64.01 | 47.36 | 0.3468 | 60.27 | 59.76 | 0.0106 |
| DPP-4 inhibitors, % | 11.45 | 3.92 | 0.2366 | 8.37 | 7.92 | 0.0143 |
| SGLT2 inhibitors, % | 2.58 | 0.53 | 0.1296 | 1.59 | 1.33 | 0.0027 |

Values are means for continuous variables and percentages for categorical variables. COPD: chronic obstructive pulmonary disease. SMD: standardized mean difference.

After propensity score matching, covariate balance improved substantially across all domains. Absolute SMD values were below 0.10 for all covariates after matching, indicating adequate covariate balance. Overall, the matching procedure substantially improved the comparability of demographic, clinical, laboratory, and treatment characteristics between statin users and non-users.

In addition, Supplementary Figure S6 presents a Love plot summarizing the absolute standardized mean differences for all covariates before and after matching. The plot reinforces the findings reported in Table 4, showing that substantial baseline imbalances, particularly in dyslipidemia, LDL cholesterol, type 2 diabetes, BMI, and the use of cardiometabolic medications, were markedly reduced following propensity score matching. After matching, absolute SMD values were below the conventional threshold of 0.10 for all covariates, indicating good overall balance between the matched groups. Overall, the matched cohorts showed good covariate balance across demographic, clinical, laboratory, and treatment domains.

Following propensity-score matching, we conducted a competing-risks analysis on the matched cohorts, treating death recorded in the CIBELES registry as the competing event. The association was slightly attenuated after matching and additional covariate adjustment but remained consistent, with statin exposure associated with a lower cumulative incidence of AMI in the presence of competing mortality (sHR 0.852; 95% CI 0.738–0.983) (Table 5), supporting the robustness of the primary finding.

**Table 5.**
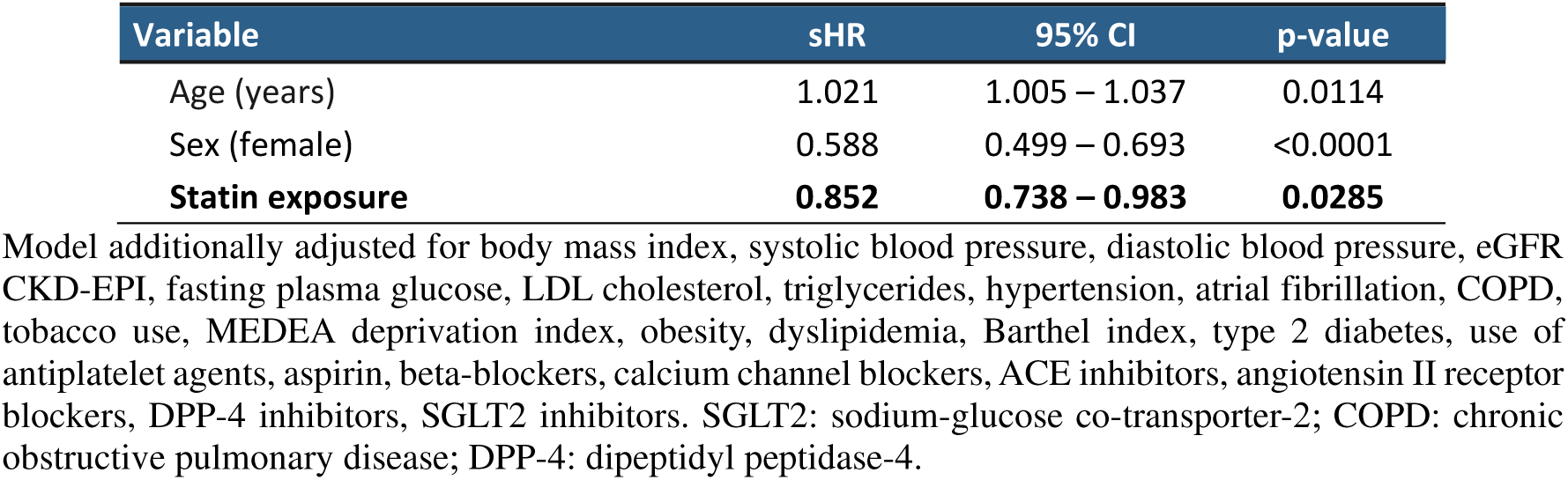
Fine–Gray competing-risk model for incident acute myocardial infarction, with mortality as the competing event in the propensity-score–matched cohort.

| Variable | sHR | 95% CI | p-value |
| --- | --- | --- | --- |
| Age (years) | 1.021 | 1.005 – 1.037 | 0.0114 |
| Sex (female) | 0.588 | 0.499 – 0.693 | <0.0001 |
| **Statin exposure** | **0.852** | **0.738 – 0.983** | **0.0285** |

Model additionally adjusted for body mass index, systolic blood pressure, diastolic blood pressure, eGFR CKD-EPI, fasting plasma glucose, LDL cholesterol, triglycerides, hypertension, atrial fibrillation, COPD, tobacco use, MEDEA deprivation index, obesity, dyslipidemia, Barthel index, type 2 diabetes, use of antiplatelet agents, aspirin, beta-blockers, calcium channel blockers, ACE inhibitors, angiotensin II receptor blockers, DPP-4 inhibitors, SGLT2 inhibitors. SGLT2: sodium-glucose co-transporter-2; COPD: chronic obstructive pulmonary disease; DPP-4: dipeptidyl peptidase-4.

## Discussion

The crude incidence rate of AMI was 4.31 per 1000 person-years, a figure comparable to that reported by de Miguel-Yanes et al. (11), who observed rates of 6.19 per 1000 person-years in men and 3.27 per 1000 person-years in women among individuals aged 75 years and older. Similarly, a prospective primary care study in Tarragona (Spain) reported AMI incidence rates of 6.32 per 1000 person-years in individuals aged 70–79 years and 6.90 per 1000 person-years in those aged ≥80 years; among participants without prior ischemic heart disease, the corresponding rates were 4.33 and 4.63 per 1000 person-years, respectively (12). These figures align closely with the rates observed in our cohort, reinforcing the external validity of our findings.

In this large population-based cohort of adults aged ≥75 years without previous cardiovascular disease, baseline statin exposure was associated with an approximately 20% lower hazard of incident acute myocardial infarction over five years. The consistency of this association across multivariable adjustment, propensity-score matching, and competing-risk analyses supports the internal coherence of the findings and provides relevant real-world observational evidence in a population that remains underrepresented in randomized trials. By focusing exclusively on acute myocardial infarction, a mechanistically coherent coronary endpoint, these results offer complementary insight into the potential relevance of the observed association in very old adults, although residual confounding cannot be excluded.

Among randomized trials, PROSPER (13) remains the only study specifically designed to evaluate statin therapy in older adults. Although pravastatin significantly reduced the primary composite endpoint among participants with established cardiovascular disease (HR 0.78; 95% CI, 0.66–0.93), no statistically significant benefit was demonstrated in the primary-prevention subgroup (HR 0.94; 95% CI, 0.77–1.15). Importantly, coronary outcomes were reported only for the overall study population and not separately for participants without previous cardiovascular disease, limiting conclusions regarding coronary prevention in this subgroup. Likewise, the long-term post-PROSPER (14) follow-up showed a lower risk of coronary heart disease death or myocardial infarction in the Scottish cohort (HR 0.79; 95% CI, 0.66–0.96), but subgroup-specific estimates for primary prevention were not available.

Evidence from other randomized trials is similarly limited. In JUPITER (15) rosuvastatin reduced myocardial infarction among older participants with high-sensitivity C-reactive protein, suggesting that anti-inflammatory and plaque-stabilizing effects may be particularly important in this age group. In HOPE-3, rosuvastatin reduced adjudicated myocardial infarction when all coronary events were considered, although significance was lost when analyses were restricted to hospitalized infarctions because of the small number of events.

These findings are broadly consistent with the Cholesterol Treatment Trialists’ Collaboration individual-participant meta-analysis (16), which found no statistically significant reduction in major vascular events among adults aged ≥75 years without established vascular disease (RR per 1 mmol/L LDL-cholesterol reduction, 0.92; 95% CI, 0.73–1.16). Rather than indicating treatment inefficacy, this result likely reflects the limited representation of very old adults and the relatively small number of cardiovascular events available for analysis in primary-prevention trials.

Accordingly, contemporary European and American guidelines emphasize individualized decision-making rather than routine statin initiation in adults older than 75 years without established cardiovascular disease. The ongoing STAREE trial (17) is expected to provide the first adequately powered randomized evidence specifically addressing this population.

Observational studies have produced heterogeneous findings. Ramos et al. (18) reported no significant association between statin use and cardiovascular outcomes among adults aged ≥75 years without diabetes, whereas statin use was associated with lower cardiovascular event rates in those with diabetes. Similarly, the Cardiovascular Health Study (19) suggested a lower incidence of myocardial infarction among statin users aged ≥74 years, although estimates were imprecise because of the limited number of events (HR 0.42; 95% CI, 0.15–1.14).

Similarly, a propensity score–matched cohort of physicians aged >76 years found no reduction in composite cardiovascular events (HR 1.05; 95% CI 0.77–1.45), although interpretation is limited by the use of a heterogeneous composite endpoint including myocardial infarction, stroke, and coronary revascularization (20).

Compared with several previous observational studies, our analysis focused exclusively on acute myocardial infarction rather than composite cardiovascular outcomes, incorporated competing-risk methods appropriate for an elderly population and included more than 170,000 individuals. These methodological features may explain why a consistent association with coronary protection was observed despite the uncertainty reported in previous studies.

The observed reduction in acute myocardial infarction is biologically plausible. Statins influence several pathways relevant to coronary atherosclerosis, including LDL-cholesterol lowering, plaque stabilization, improvement of endothelial function, and attenuation of vascular inflammation. These mechanisms may reduce plaque vulnerability and the likelihood of coronary thrombosis, and may be particularly pertinent in older adults, in whom vascular aging and chronic low-grade inflammation accelerate atherosclerotic progression. Although causality cannot be inferred from an observational study, the concordance between these well-established biological mechanisms and the present findings supports the plausibility of the observed association.

Although the relative reduction in AMI risk was approximately 20%, the absolute difference in AMI risk was modest (0.30%), corresponding to an estimated observational number needed to treat of 338 over 5 years (95% CI 222–708), interpreted as a mathematical translation of the absolute risk difference. This distinction between relative and absolute measures is particularly relevant in primary prevention among very old adults, as the absolute difference depends strongly on baseline cardiovascular risk and competing mortality. Thus, while the observed association indicates lower AMI incidence among statin users, the magnitude of the absolute difference is likely to be greatest in older adults with sufficiently high cardiovascular risk and life expectancy.

### Strengths

This study has several strengths. First, it included a large, contemporary, population-based cohort of 174,014 community-dwelling adults aged ≥75 years, a population that remains underrepresented in randomized trials of primary prevention. The size and population-based nature of the cohort provided substantial statistical power to evaluate incident AMI in a clinically relevant group of very old adults without prior cardiovascular disease.

Second, the study integrated data from multiple complementary sources, including primary-care electronic medical records, hospital discharge records, laboratory information systems, and pharmacy dispensing records. This integration enabled the assessment of diagnoses, cardiovascular risk factors, functional status, laboratory measurements, socioeconomic context, medication exposure, and mortality within a common analytical framework.

Third, the landmark-based design established a clear temporal sequence between exposure assessment and outcome follow-up. Statin exposure was classified using dispensing records from 2018–2019, before prospective follow-up for incident AMI began on 1 January 2020. This fixed baseline exposure strategy avoided exposure classification during outcome follow-up and provided a consistent framework for comparing participants, while subsequent treatment changes were handled in accordance with the predefined baseline classification.

Fourth, the analysis incorporated several clinically relevant domains that may influence treatment selection and cardiovascular risk in older adults, including functional status, multimorbidity, laboratory measurements, concomitant medication use, and area-level socioeconomic deprivation. The use of multiple imputation allowed the analysis to retain participants despite incomplete covariate information, while the inclusion of propensity-score matching provided a complementary approach to address measured baseline differences between exposure groups.

Fifth, the study used a previously validated coding approach to identify incident AMI from primary-care and hospital data and evaluated the outcome using both multivariable Cox regression and competing-risk methods. Accounting for all-cause mortality as a competing event was particularly relevant in this age group, in which death may preclude the occurrence or ascertainment of a subsequent AMI. The consistency of the association across the primary and sensitivity analyses supports the internal coherence of the observed findings, although it does not eliminate the possibility of residual confounding or establish causality.

### Limitations

This study has several limitations. First, residual confounding cannot be fully excluded despite extensive adjustment for demographic, clinical, functional, laboratory, pharmacological, and socioeconomic variables. Unmeasured factors, including frailty trajectories, cognitive and nutritional status, clinician prescribing preferences, treatment goals, health-seeking behavior, and adherence-related characteristics, may have influenced both statin initiation and AMI risk. Several covariates had substantial missingness, and although multiple imputation was performed under a missing-at-random assumption, this assumption cannot be verified, and differential patterns of missingness may remain.

Second, statin exposure was defined using a fixed baseline strategy analogous to an observational intention-to-treat approach. This avoided time-dependent exposure classification but did not account for treatment changes after baseline, potentially attenuating the observed association. Because eligibility required survival until the landmark date, healthy-survivor bias cannot be fully excluded, and its direction and magnitude are uncertain. Nonetheless, exposure status was fully defined before follow-up began, reducing the risk of immortal-time bias. Information on statin intensity was not available, preventing evaluation of whether higher-intensity regimens confer greater protection or whether treatment effects differ across dosing strategies commonly used in older adults.

Third, AMI was identified from routinely collected primary-care and hospital data rather than through individual adjudication. Although validated diagnostic codes were used, misclassification remains possible, particularly for atypical, silent, or non-hospitalized events. Cause-specific mortality was not available, limiting the ability to distinguish cardiovascular from non-cardiovascular deaths. The Fine– Gray competing-risk model therefore estimates associations with cumulative incidence of AMI rather than causal effects.

Finally, the analysis focused exclusively on AMI and did not evaluate other clinically relevant outcomes such as ischemic or hemorrhagic stroke, heart failure, cardiovascular mortality, adverse drug effects, functional status, or treatment burden. The study was conducted in a single European region with universal healthcare coverage and specific prescribing patterns, which may limit generalizability to other settings. Replication in independent cohorts would strengthen external validity and help determine whether similar associations are observed across diverse clinical and sociodemographic contexts.

### Conclusions

In this large, population-based cohort of community-dwelling adults aged ≥75 years without prior cardiovascular disease, baseline statin exposure was associated with a lower hazard of acute myocardial infarction over a median follow-up of 5 years, with consistent findings across multiple analytical approaches. Although the relative reduction in hazard was approximately 20%, the absolute difference in AMI risk was modest (0.30%), corresponding to an estimated observational number needed to treat of 338 over 5 years, interpreted as a mathematical translation of the absolute risk difference. These results highlight that the magnitude of the absolute difference in AMI incidence is likely to vary according to baseline cardiovascular risk and life expectancy. Accordingly, chronological age alone should not preclude consideration of statin therapy, but treatment decisions should remain individualized and incorporate frailty, functional status, comorbidity, treatment burden, life expectancy, and patient preferences. Further randomized evidence, including from STAREE, is needed to determine whether these observational associations translate into clinically meaningful differences in outcomes.

## Supporting information

STROBE Checklist

## Author Contributions

Conceptualization, M.A.S-F.; Methodology, J.C-V. and M.A.S-F.; Formal analysis, O.A-dC and M.A.S-F.; Data curation, J.C-V., O.A-dC, F.J.S-S, and M.B-D; Validation, J.M, C.L, F.J.S-R, P.V-P, and A.I.G-G; Writing, original draft preparation, M.A.S-F; Writing, review and editing, J.C-V, O.A-dC, M.B-D, P.V-P, A.I.G-G, J.M., and C.L.; Funding acquisition, A.I.G-G.; Supervision, J.C-V., M.B-D and A.I.G-G. All authors have read and agreed to the published version of the manuscript.

## Funding

This project (“HealthData@MAD-R&I^®^”, code TSI-100121-2024-79) is funded by the Ministry for Digital Transformation and Public Service and by the European Union through financial resources derived from the European Recovery Instrument (“Next Generation EU”), within the framework of the Recovery, Transformation, and Resilience Plan.

However, the views and opinions expressed are solely those of the authors of this manuscript and do not necessarily reflect those of the European Union or the European Commission. Neither the European Union nor the European Commission can be held responsible for them.

## Institutional Review Board Statement

The study was conducted in accordance with the Declaration of Helsinki and the General Data Protection Regulation (GDPR). It was approved by the Regional Research Ethics Committee on Medicines of Madrid (CEIM-R) on January 13, 2025 (protocol code 01_sept_2024).

## Informed Consent Statement

The Ethics Committee did not require informed consent because the research was performed with secondary data.

## Data Availability Statement

Data sharing is currently restricted while the HealthData@MAD-R&I^®^ infrastructure remains under development. However, selected non-sensitive materials will be deposited in the RIMASalud repository. Upon completion of the project, additional non-sensitive materials, including technical documentation and example scripts, will also be deposited in RIMASalud for open access. Requests for access to restricted datasets may be submitted through the governance procedures of the Madrid Regional Health System (SERMAS) once the Secure Processing Environments become operational.

## Acknowledgments

The authors acknowledge the use of AI-based tools, including Perplexity (version 3; developed by Perplexity AI Inc.) and Claude Sonnet 5 (Anthropic), to assist with code review, error checking, and language editing during manuscript preparation. These tools were used solely as supportive resources. All study design decisions, statistical analyses, interpretation of results, and conclusions were conducted and verified by the authors, who take full responsibility for the content of the manuscript.

