## Supplementary Material for "Baseline Statin Exposure and Incident Acute Myocardial Infarction in Adults Aged ≥75 Years Without Prior Cardiovascular Disease: A Population-Based Cohort Study"

**Supplementary Figure S1. Kaplan-Meier curve according to statin use in men.**

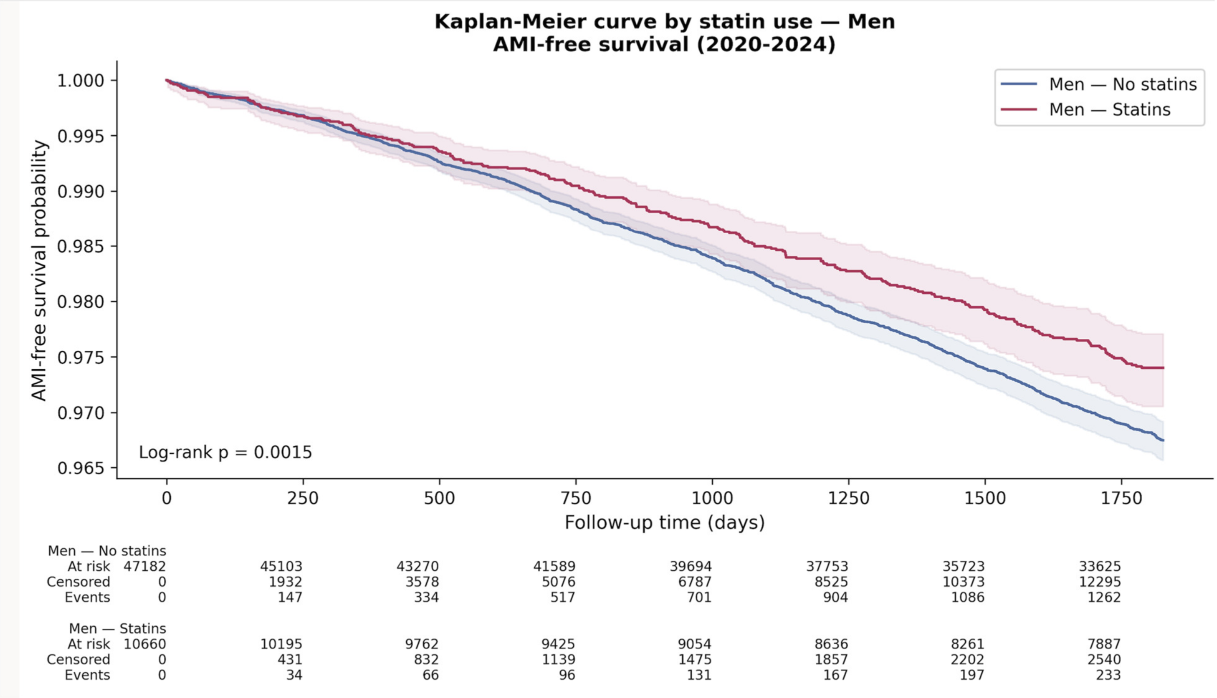

**Supplementary Figure S2. Kaplan-Meier curve according to statin use in women.**

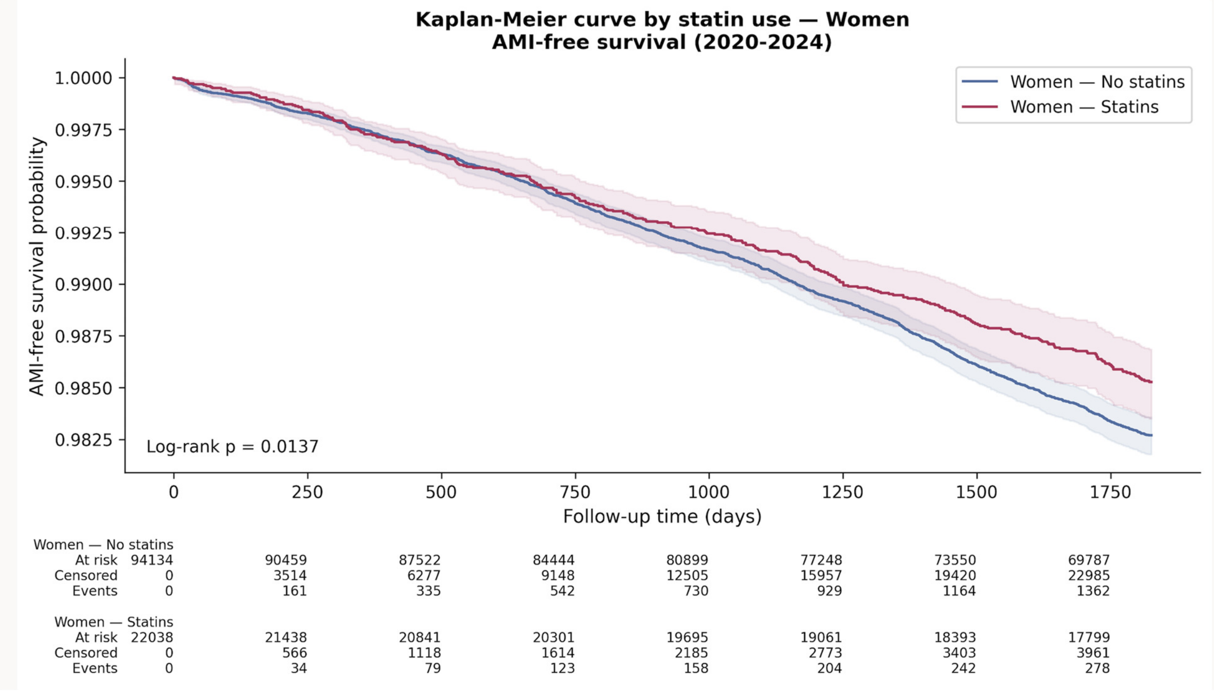

**Supplementary Table S1. Proportion of missing values.**

| Variable | % |
|---|---|
| Waist circumference | 93.5 |
| Microalbuminuria | 72.6 |
| Lawton-Brody (IADL category) | 71.2 |
| Barthel Index category | 51.1 |
| eGFR (CKD-EPI) | 48.0 |
| BMI | 46.6 |
| Number of LDL cholesterol measurements | 35.5 |
| LDL cholesterol (last value 2018-2019) | 35.5 |
| Triglycerides (last value 2018-2019) | 32.5 |
| Fasting plasma glucose (last value 2018-2019) | 31.3 |
| Diastolic blood pressure (last value 2018-2019) | 26.5 |
| Systolic blood pressure (last value 2018-2019) | 26.4 |
| Number of Systolic blood pressure measurements | 26.4 |
| Deprivation index | 4.7 |

**Supplementary Figure S3. Distribution of Original and Imputed Data for Clinical Variables (20-Dataset Multiple Imputation)**

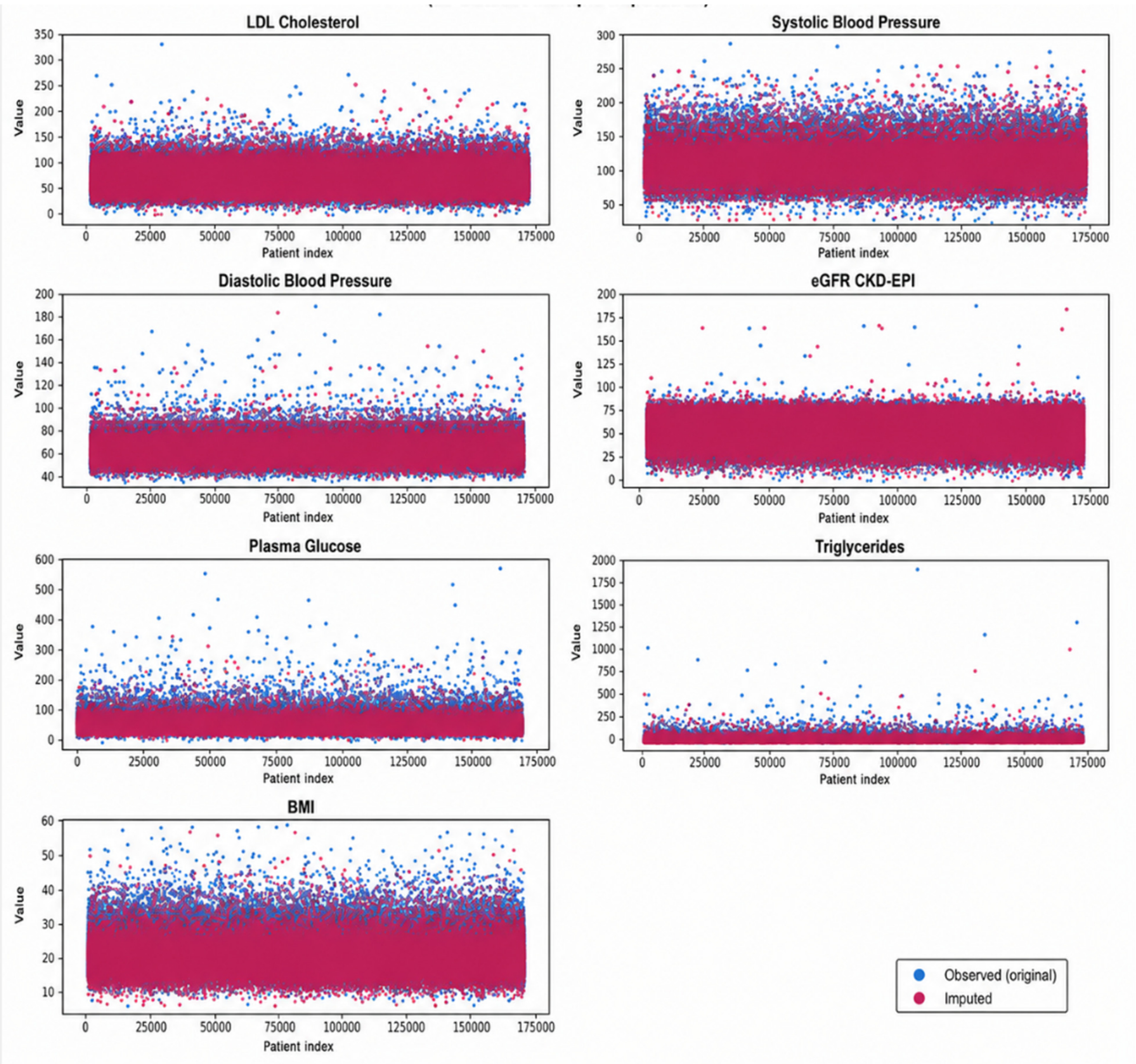

**Supplementary Figure S4. Density Plots Comparing Original and Imputed Distributions Across 20 Multiple-Imputation Datasets**

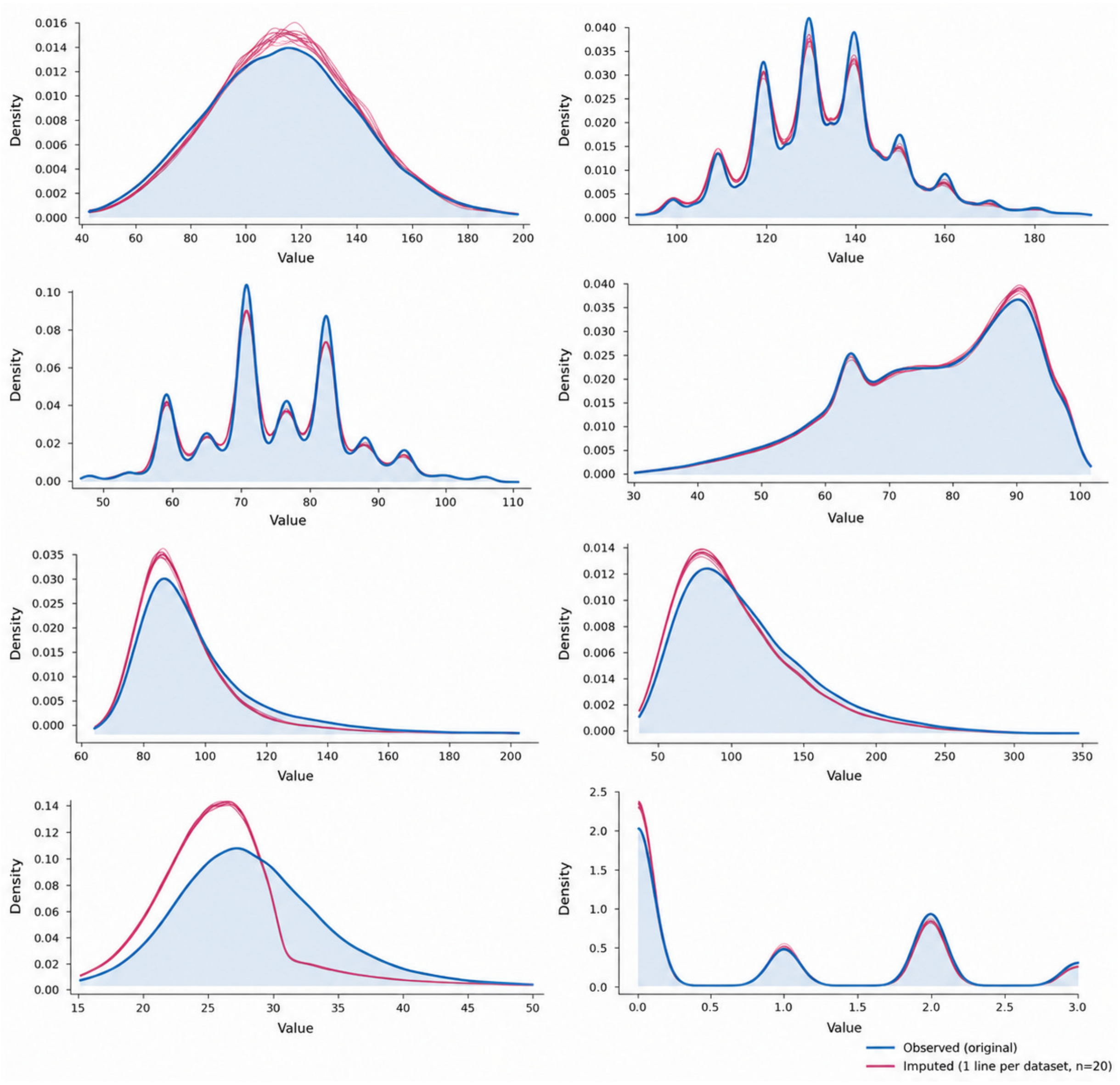

**Supplementary Figure S5. Diagnostic plots for the Cox proportional hazards model: Martingale residuals versus follow-up time and deviance residuals.**

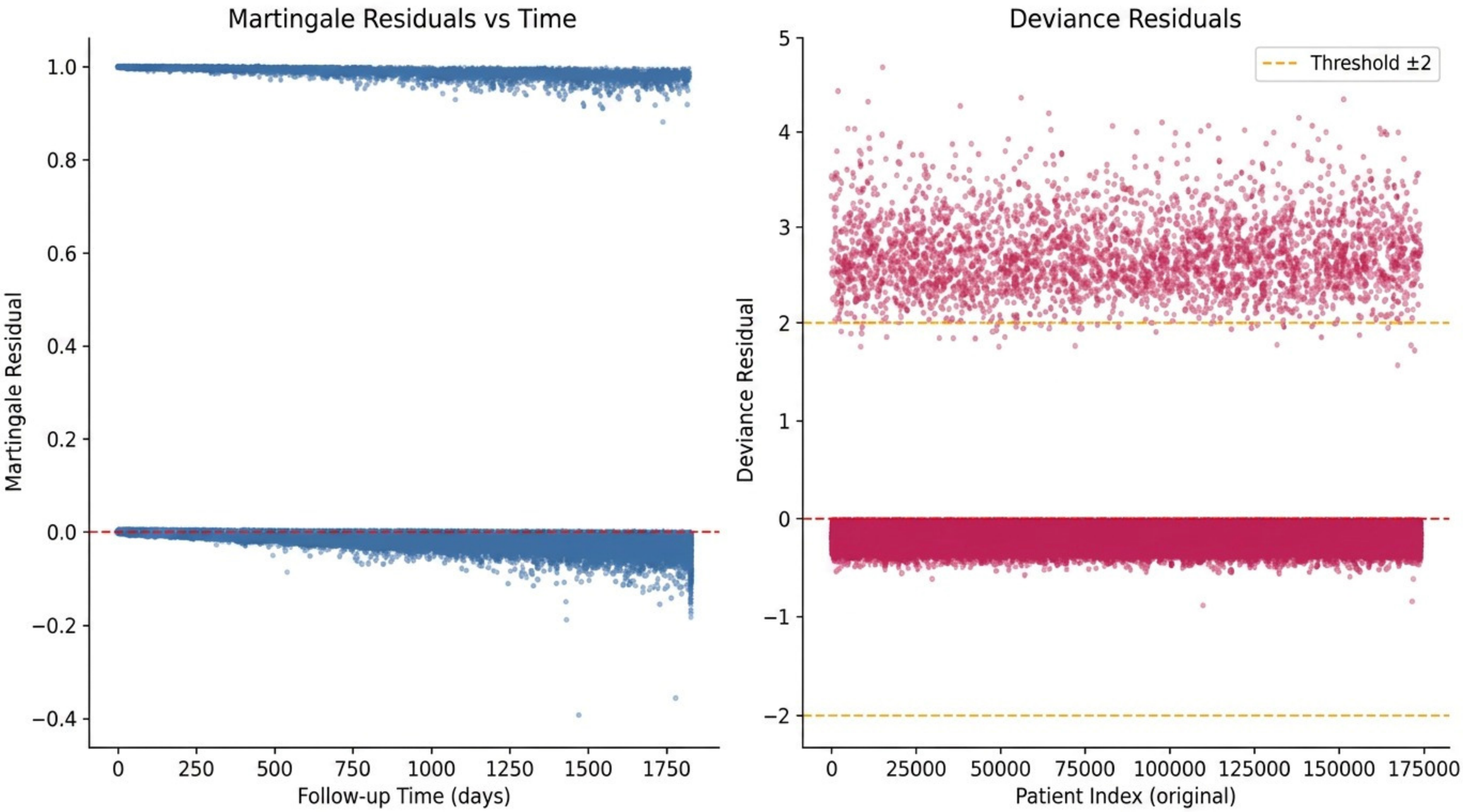

**Supplementary Figure S6. Love Plot**

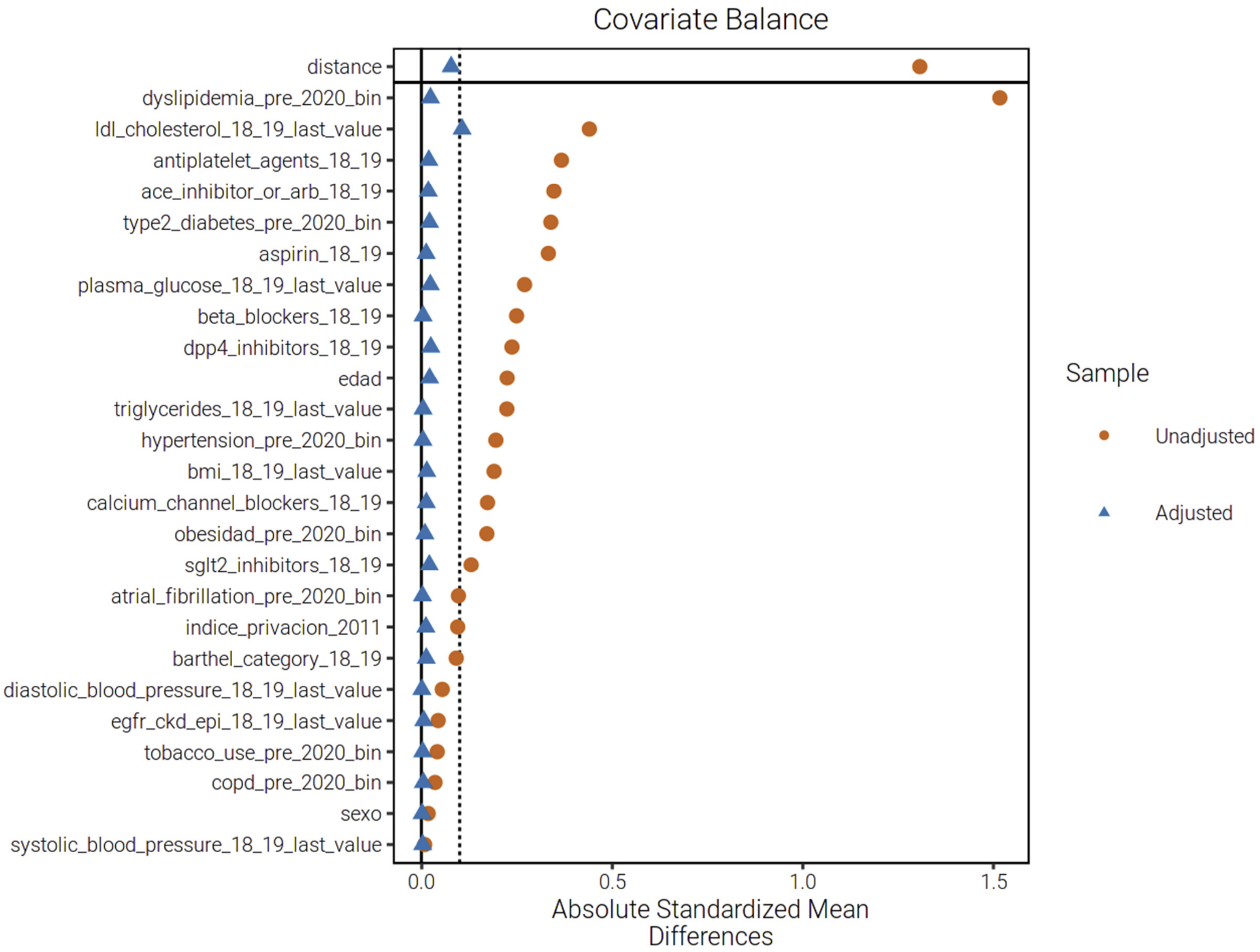
