## Supplementary material for "Baseline Statin Exposure and Incident Acute Myocardial Infarction in Adults Aged ≥75 Years Without Prior Cardiovascular Disease: A Population-Based Cohort Study": STROBE Checklist

### STROBE Cohort Checklist – Completed for medRxiv submission

This working checklist maps the STROBE cohort reporting items to the final submission PDF. It is intended as submission/support documentation; it does not modify the manuscript.

| Item | Section | Reporting element | Location in final PDF |
|---|---|---|---|
| 1a | Title/Abstract | Study design identified with a standard design term. | pp. 1–2 (title/abstract) |
| 1b | Title/Abstract | Abstract provides a balanced summary of methods and findings. | pp. 1–2 |
| 2 | Introduction | Scientific background and rationale. | pp. 2–3 |
| 3 | Introduction | Study objective stated explicitly. | p. 3 |
| 4 | Methods | Key elements of the cohort design presented early. | pp. 3–4 |
| 5 | Methods | Setting, region, landmark date, exposure period and follow-up dates. | pp. 3–4, 7 |
| 6a | Methods | Eligibility criteria, participant selection and follow-up. | pp. 4, 7 |
| 6b | Methods | Matching approach and matched group sizes. | pp. 8, 17–18 |
| 7 | Methods | Outcome, exposure and covariates defined. | pp. 4–5 |
| 8 | Methods | Sources and measurement/ascertainment methods described. | pp. 4–6 |
| 9 | Methods | Approaches relevant to bias/confounding described, including landmark design, multivariable adjustment, PSM and competing risks. | pp. 4, 6–8 |
| 10 | Methods | Study size derives from the eligible regional population; no formal sample-size calculation is reported. | pp. 4, 8 |
| 11 | Methods | Handling of quantitative variables and prespecified measurements described. | pp. 5–7 |
| 12a | Methods | Statistical methods, including confounder adjustment. | pp. 6–8 |
| 12b | Methods | Subgroup/interaction methods, where applicable. Sex-stratified incidence described. | pp. 7, 13 |
| 12c | Methods | Missing-data methods. | p. 6 |
| 12d | Methods | Loss to follow-up/censoring handled in follow-up definition. | p. 7 |
| 12e | Methods | Sensitivity analyses described. | pp. 7–8 |
| 13a | Results | Participant flow and final cohort reported. | p. 9; Figure 1 |
| 13b | Results | Reasons for exclusion represented in study flow. | p. 9; Figure 1 |
| 13c | Results | Flow diagram provided. | p. 9; Figure 1 |
| 14a | Results | Baseline demographic/clinical characteristics reported. | pp. 10–12; Table 1 |
| 14b | Results | Missingness reported. | pp. 6, 11–12; Supplementary Table S1 |
| 14c | Results | Follow-up duration summarized. | pp. 2,22 |
| 15 | Results | Outcome events and incidence measures reported. | p. 13 |

| | | | |
|---|---|---|---|
| 16a | Results | Unadjusted and adjusted estimates with 95% CIs reported. | pp. 13, 15–18 |
| 16b | Results | Not applicable: the primary exposure (statin use) was binary; no continuous exposure variable was categorized. | N/A |
| 16c | Results | Absolute risk translated to an observational NNT with explicit interpretation. | pp. 2, 13, 20, 22 |
| 17 | Results | Other/sensitivity analyses reported (PSM and competing-risk analyses). | pp. 16–18 |
| 18 | Discussion | Key results summarized with reference to objectives. | pp. 18–20 |
| 19 | Discussion | Study limitations and potential sources of bias/imprecision discussed. | pp. 21–22 |
| 20 | Discussion | Overall interpretation is cautious and considers observational design. | pp. 18–22 |
| 21 | Discussion | Generalizability/external validity discussed. | pp. 18, 22 |
| 22 | Other information | Funding source and role/disclaimer reported. | p. 22 |

Reporting guideline: STROBE (cohort studies). The official STROBE checklist is available from the STROBE Initiative/EQUATOR Network.
